# Walking Gaze Behavior After a Stroke: More Than Meets the Eye

**DOI:** 10.1101/2025.05.08.25326839

**Authors:** Yogev Koren, Noy Goldhamer, Shilo Kramer, Lior Shmuelof

## Abstract

A common clinical impression is that individuals with stroke tend to gaze downward while walking—focusing on the walking surface a short distance ahead for extended periods. However, this impression has not been formally verified, and thus, whether this is a true phenomenon—and, if so, what drives it—remains unknown.

In this observational study, we examined the spatial and temporal aspects of walking gaze behavior in individuals with stroke and compared them to those of healthy controls. Our results indicate that individuals with stroke exhibit a greater tendency to gaze downward while walking compared to healthy controls.

While the shorter look-ahead distances observed may be attributed to slower walking speeds, the prolonged duration of downward gazing (DWG) cannot be explained by speed alone. Instead, both the short look-ahead distance and prolonged DWG duration were associated with anxiety—particularly fear of falling—as well as impaired balance and gait control. Importantly, while participants reported consciously monitoring their stepping, this tendency was unrelated to DWG, suggesting that downward gaze is unlikely to serve this specific purpose.

These findings suggest that anxiety related to walking instability may underlie both the slower walking speeds and the tendency for DWG. Having established the presence of the DWG phenomenon, we propose further investigation into its potential utility as a novel indicator of individuals’ self-assessed deficits in reactive and/or proactive balance and gait control.

## Background

A common clinical observation is that unstable walkers tend to gaze down while walking. That is, they look at the walking surface, a short distance ahead, for extended periods regardless of whether foot-placement accuracy is required.

Importantly, while induced walking instability was shown to cause downward gazing [1] our clinical impression in pathological populations has little, if any, quantitative support. Thus, it is currently unclear whether this impression is true, and if so, whether it is driven by walking instability.

Stroke is a medical condition in which disturbance to blood flow to the brain leads to cell death. Stroke often leads to motor, sensory, and cognitive impairments, resulting in walking instability. Persons with stroke (PwS) are unstable in the sense that they are prone to falling [2–5], even years following the event [see review in [6]]. Therefore, if our clinical impression is of a true phenomenon, it would be expected that PwS gaze down more than controls while walking and that this propensity be associated with walking instability.

While there is no doubt that visual input is constantly used during walking [see several reviews in [7–10]], to understand the visual control of human locomotion, it is critical “to determine when, where, what and how different visual information is acquired” [9]. In line with Patla’s suggestion, over the years many researchers [11–21] have used eye-tracking technology to determine the spatiotemporal pattern of walking gaze behavior, mostly [but not exclusively, e.g.,[22–25]] targeting scenarios in which accuracy of the foot’s trajectory is important. Others [26–33] have used a complementary approach and manipulated the visual input to determine whether and how this manipulation affects stepping and walking.

That said, walking gaze behavior is not just about acquiring visual input [17, 34], and has been suggested to be driven by a whole-body coordination pattern [35]. From both perspectives, deviation from a typical gaze pattern might offer a glimpse into impaired control in pathological conditions. With that in mind, in this investigation we provide a qualitative and a quantitative description of the walking gaze behavior of PwS and compare it to that of healthy controls. Furthermore, following the hypothesis that gaze behavior in PwS is the consequence of poor control and instability, we examine the associations between the observed spatial and temporal gaze patterns and the steadiness of posture and gait, fear of falling and the tendency for conscious movement processing (CMP).

## Results

Participants performed the 10-meter walk test, along a ∼15m course, while wearing a mobile eye-tracker (see details in the Methods section). Participants were PwS (106 trials), aged-matched controls (30 trials) and younger controls (24 trials), instructed to walk at their preferred speed. Descriptive statistics of the demographics of these participants are summarized in Table 1.

**Table 1.** Descriptive statistics (median [inter-quartile range])

|  | Younger (N=12) | Older (N=15) | Stroke (N=31) |
| --- | --- | --- | --- |
| Age<sub>yrs</sub> | 28 [25-36] | 66 [63-73] | 69 [62-76] |
| Height<sub>cm</sub> | 168 [161-175] | 163 [153-174] | 167 [162-171] |
| Weight<sub>kg</sub> | 79 [51-79] | 68 [60-79] | 76 [68-85] |
| Sex<sub>F/M</sub> | 10/2 | 11/4 | 5/26 |
| Paretic side<sub>L/R</sub> |  |  | 9/22 |

The experimental setup, and the main gaze outcome measures are presented graphically and briefly explained in Figure 1. A detailed explanation appears in the Methods section.

**Figure 1.**
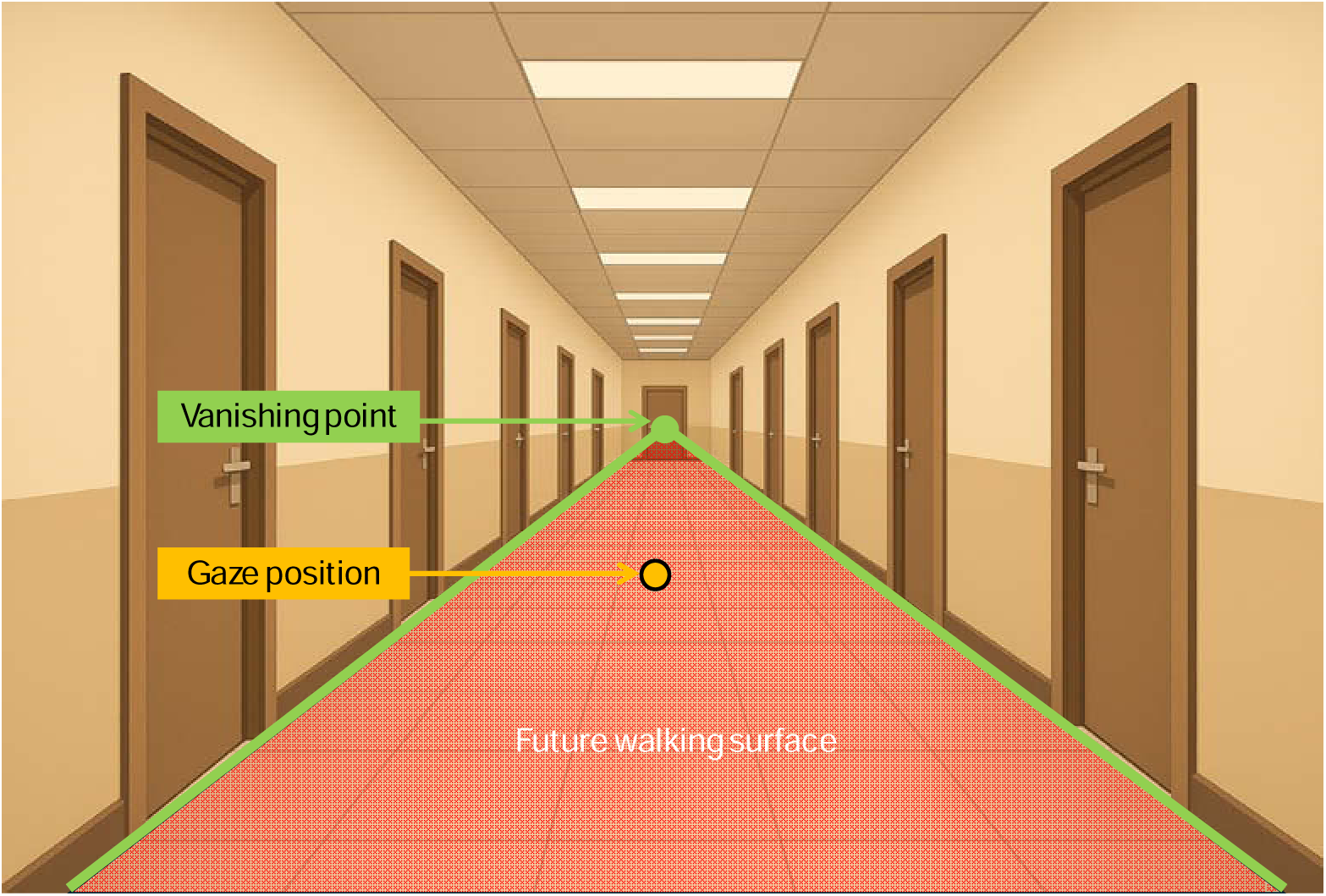
Illustration of the experimental setup. Participants wearing a mobile eye-tracker walked back and forth along a corridor. Using image processing, we identified the floor-lines (highlighted in green) and their intersection point or vanishing point (the green circle) in each frame. The area contained within this “triangle” (shaded red) is the future walking surface. Gaze positions projected onto the walking surface (shaded red) are “on surface” and are reported as a percentage of all gaze positions during the entire walk (DWG duration). Each “on surface” position has a unique distance from the observer, reported in meters (DWG distance). Analysis of additional outcome measures are explained and presented in the Supplementary Materials section.

The gaze behaviors of the younger and older controls were not different (spatially or temporally), so they were combined into a single control group (for analysis of the 3 groups, see Supplementary Materials, FigureS2). Comparison of the gaze behaviors of PwS and controls are represented graphically in Figure 2a. Generally, gaze positions were classified as either ‘on’ or ‘off’ the walking surface (see Figure 1 for explanation). For each ‘on surface’ position, a look-ahead distance was calculated (i.e., the distance from the observer). In Figure 2, gaze distances were binned into 0.5-m intervals. For each group, we calculated the mean probability of each bin.

**Figure 2.**
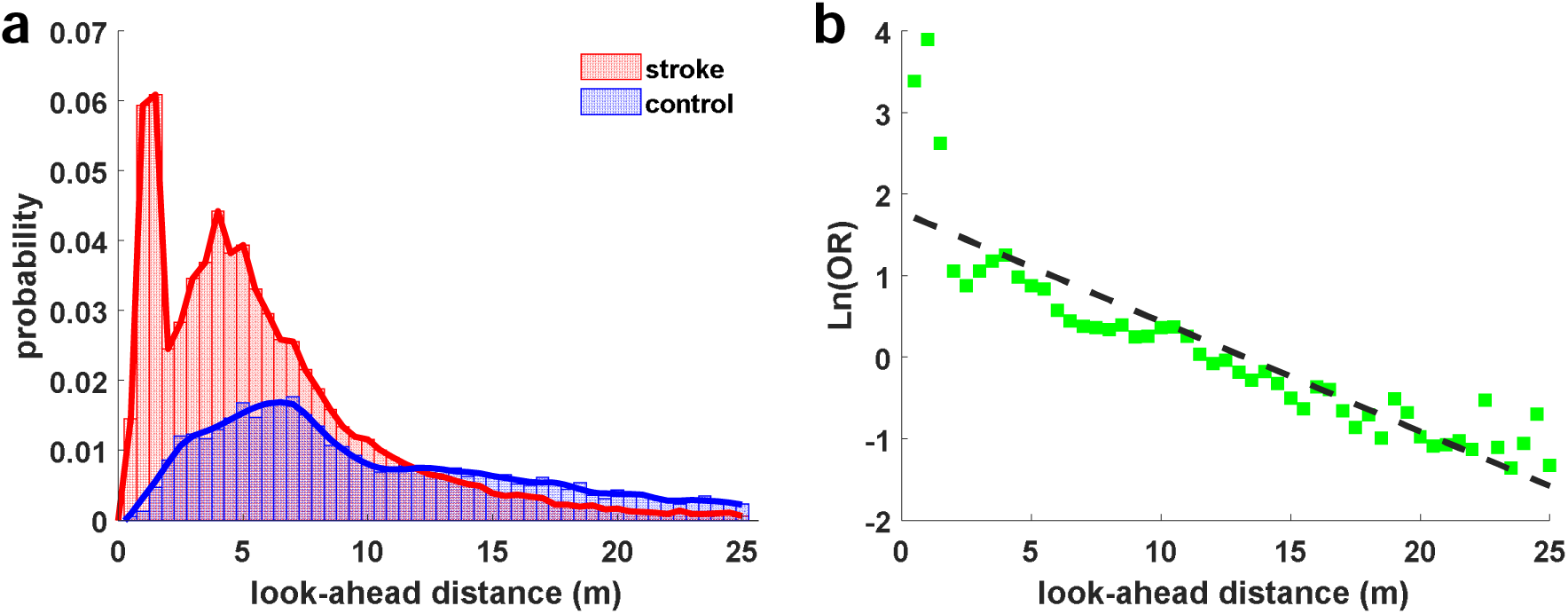
**Panel a** presents the mean probability of the look-ahead distances (up to 25m) by group. “Probability” is the ratio of the number of samples observed for a certain bin and the total number of samples in the walk. These values are averaged for each group. The width of each bin is 0.5m, and the area under the curve represents the percentage of time participants looked onto the walking surface. **Panel b** presents the odds-ratio (log-transformed values) of the probability between PwS and controls. These ratios are plotted against the look-ahead distance and the dotted line denotes the linear predictor of the regression.

Here, probability is the number of samples observed for a certain bin divided by the total number of samples during the entire walk (both On and Off surface). The figure shows that PwS looked for longer at the walking surface (represented by the area under the curve) and did so by increasing the percentage of time spent looking at the immediate surface, but also at greater distances of up to ∼7m ahead. The figure also shows two peaks for PwS, the first at ∼ 1.5m ahead and the second at ∼4m ahead.

For controls, a single peak at ∼7m ahead was observed.

The impression from Figure 2a is that most of the difference between PwS and controls is in the short/medium look-ahead range (roughly up to 7m ahead). This might indicate that PwS change their walking gaze behavior temporally, to increase only the time spent looking up to ∼7m ahead, while maintaining the time spent looking further ahead. To investigate the differences between groups throughout the range, we calculated the odds-ratio (PwS/controls) of the probabilities for each bin and tested the relation between these ratios and the look-ahead distance. We reason that if the differences were confined to a certain look-ahead range, then the odds-ratio would asymptote to 1 at greater look-ahead distances. The results of this analysis are presented graphically in Figure 2b. In this figure, it is notable that the odds-ratio is linearly related to the look-ahead distance (p=0.00, R^2^=0.81).

Specifically, the odds of looking at the immediate surface is ∼4 times greater for a PWS than for a healthy person, but these odds decrease as the look-ahead distance increases. This linear decrease indicates that odds-ratio does not asymptote to 1 (zero in the figure, as values were log-transformed), but instead continues to decrease throughout the range, and at 25m ahead, controls are ∼4 times more likely to gaze than PwS. This analysis indicates that the whole distribution curve, rather than just a portion, is affected.

To test the differences between groups, we compared gaze duration and distance between them. We also binned gaze into 3 distance categories [see Methods and [1]] and compared the duration in each bin between groups. (Comparisons of additional parameters are reported in the Supplementary Materials TableS1.) These comparisons are summarized in Table 2. Briefly, overall, PwS gazed for longer at the walking surface, and to a shorter distance ahead than did controls, indicating spatial and temporal differences. At the same time, PwS spent less time than controls looking at distances greater than 10m ahead.

**Table 2.**
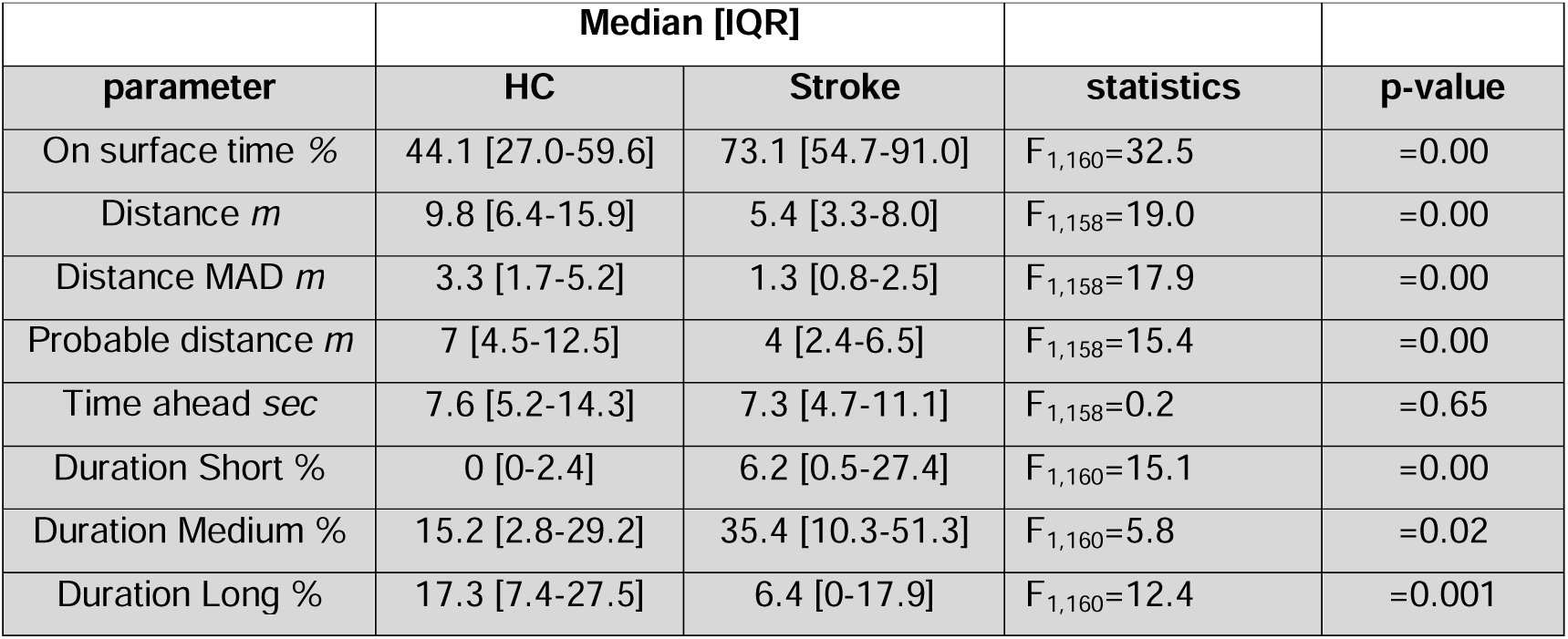
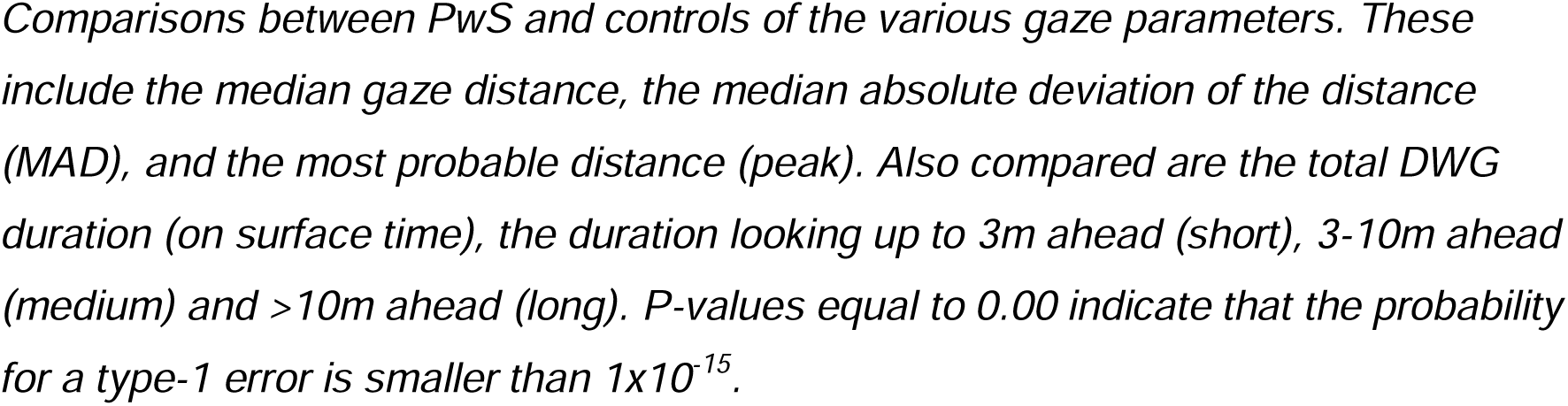
Comparisons of gaze parameters between groups.

|  | **Median [IQR]** |  |  |  |
| --- | --- | --- | --- | --- |
| **parameter** | **HC** | **Stroke** | **statistics** | **p-value** |
| On surface time % | 44.1 [27.0-59.6] | 73.1 [54.7-91.0] | $F_{1,160}=32.5$ | =0.00 |
| Distance *m* | 9.8 [6.4-15.9] | 5.4 [3.3-8.0] | $F_{1,158}=19.0$ | =0.00 |
| Distance MAD *m* | 3.3 [1.7-5.2] | 1.3 [0.8-2.5] | $F_{1,158}=17.9$ | =0.00 |
| Probable distance *m* | 7 [4.5-12.5] | 4 [2.4-6.5] | $F_{1,158}=15.4$ | =0.00 |
| Time ahead sec | 7.6 [5.2-14.3] | 7.3 [4.7-11.1] | $F_{1,158}=0.2$ | =0.65 |
| Duration Short % | 0 [0-2.4] | 6.2 [0.5-27.4] | $F_{1,160}=15.1$ | =0.00 |
| Duration Medium % | 15.2 [2.8-29.2] | 35.4 [10.3-51.3] | $F_{1,160}=5.8$ | =0.02 |
| Duration Long % | 17.3 [7.4-27.5] | 6.4 [0-17.9] | $F_{1,160}=12.4$ | =0.001 |

*Comparisons between PwS and controls of the various gaze parameters. These include the median gaze distance, the median absolute deviation of the distance (MAD), and the most probable distance (peak). Also compared are the total DWG duration (on surface time), the duration looking up to 3m ahead (short), 3-10m ahead (medium) and >10m ahead (long). P-values equal to 0.00 indicate that the probability for a type-1 error is smaller than $1 \times 10^{-15}$ .*

Our clinical impression statement defines DWG in terms of duration and distance. Yet, for locomotion, visual input often serves to anticipate and plan future actions [see review in [8]]. These aspects are sensitive to the speed of locomotion. Thus, it makes sense that actions are planed ahead in time rather than space. For example, for safety reasons, drivers are taught to lag 2sec behind the car in front of them, so they have enough time to detect and react to any change. In the special case of precise stepping, this “time-ahead” strategy also seems to hold [19]. While the current task did not require precise stepping, it is still possible that the central nervous system uses a time-ahead strategy, as some general strategy. Given that PwS walked more slowly than controls (see Table 3), it is possible that the spatial differences between groups simply reflect the different walking speeds. Thus, to investigate this possibility we computed the distribution of the look-ahead times of the two groups. To do so, we simply divided each look-ahead distance by the average walking speed during the trial. These look-ahead times were binned into 0.5sec intervals, and the mean probability of each bin and group was calculated.

**Table 3.** The association between DWG and walking instability.

|  | Duration | Distance | Time | Short | Medium | Long | Peak |
| --- | --- | --- | --- | --- | --- | --- | --- |
| Psychological |  |  |  |  |  |  |  |
| CMP | 0.14 | -0.23 | -0.11 | 0.18 | 0.13 | -0.15 | -0.18 |
| Anxiety | 0.57*** | -0.43** | -0.34* | 0.43** | 0.03 | -0.32* | -0.37* |
| Standing postural sway |  |  |  |  |  |  |  |
| ML Range | 0.45*** | -0.55*** | -0.20 | 0.60*** | -0.17 | -0.55*** | -0.44*** |
| AP Range | 0.58*** | -0.53*** | -0.13 | 0.58*** | -0.05 | -0.42** | -0.45** |
| Area | 0.54*** | -0.55*** | -0.14 | 0.59*** | -0.09 | -0.49*** | -0.45** |
| Velocity | 0.44** | -0.45** | -0.08 | 0.50*** | -0.26 | -0.36** | -0.34** |
| Drs | 0.54*** | -0.53*** | -0.16 | 0.57*** | -0.18 | -0.46** | -0.43** |
| Dxs | 0.52*** | -0.50*** | -0.14 | 0.56*** | -0.21 | -0.46** | -0.41** |
| Dys | 0.52*** | -0.53*** | -0.08 | 0.57*** | -0.18 | -0.44** | -0.45** |
| Walking speed and step variability |  |  |  |  |  |  |  |
| Speed | -0.39** | 0.40** | -0.25 | -0.48*** | 0.18 | 0.25 | 0.42** |
| Time SD | 0.28* | -0.35* | 0.15 | 0.46** | 0.17 | -0.24 | -0.34* |
| Length SD | 0.13 | -0.16 | 0.07 | 0.17 | 0.00 | -0.10 | -0.16 |
| Width SD | 0.24 | -0.30* | -0.05 | 0.36** | -0.37** | -0.34* | -0.29* |
| Speed SD | -0.03 | 0.02 | 0.11 | 0.00 | 0.00 | 0.03 | 0.05 |

*The table represents a correlation coefficient matrix. Values are the Spearman correlation coefficients. For clarity, absolute values (not signed values) are coded in shades of blue, with darker shades representing larger absolute values. For an explanation of the standing sway metrics, see Methods. SD is the standard deviation of step metric. ‘\*’, ‘\*\*’ and ‘\*\*\*’ indicate $p<0.05$ , $p<0.01$ and $p<0.001$ , respectively.*

The results of this analysis are presented in Figure 3a. This figure shows a more uniform gaze probability distribution between groups, with maximal probability (peak) observed at ∼4.5sec ahead for both groups. Indeed, testing the central tendency of the look-ahead time between groups revealed a negligible difference between them (see Table 2 for details). The odds-ratio analysis (Figure 3b) for the look-ahead times revealed a quadratic pattern over the investigated look-ahead times (p=4.4×10^-6^, R^2^=0.27), again failing to asymptote to 1 (zero for the log-transformed values). These results possibly indicate that humans adjust their look-ahead distance to their walking speed, meaning that the spatial difference between groups reflects their different walking speeds. Yet, this analysis provides no direct explanation for the temporal difference between them (i.e., the time spent looking at the walking surface).

**Figure 3.**
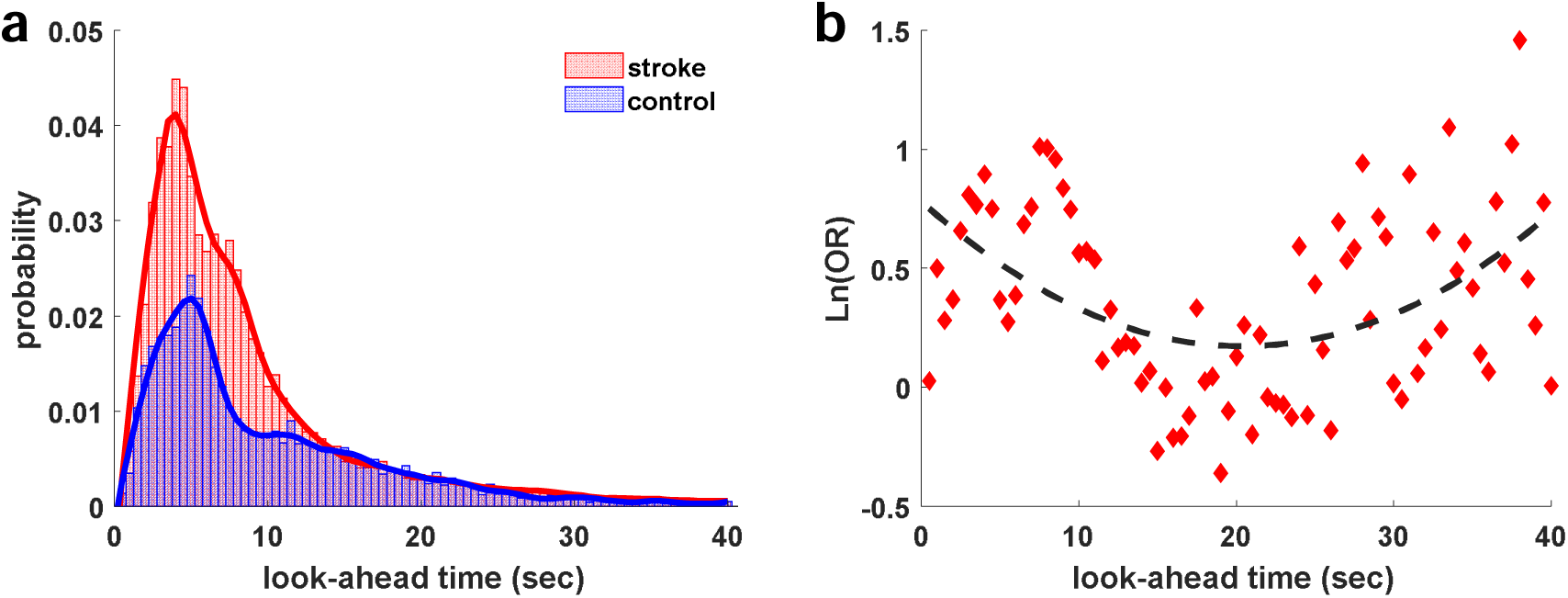
the mean probability of the look-ahead time by group. Probability is the ratio of the number of samples observed for a certain time bin to the total number of samples in the walk. These values are averaged for each group. The width of each bin is 0.5sec. In **a**, this probability is plotted against the time-ahead. In **b**, we calculated the odds-ratio of this probability between groups, and plotted these values (log-transformed) against the time-ahead.

Nonetheless, this does not mean that slow walking speed is not driving people to look longer onto the walking surface. If the difference in DWG duration between groups is a consequence of slow walking speed, then manipulating walking speed is expected to reveal this relation. Therefore, healthy control subjects were asked to walk at their preferred speed, but also at slower and faster speeds. The results of this manipulation are depicted in Figure 4. This figure shows that slow walking speed (1.2 Vs. 0.95 m/sec, p=0.00) had no effect on DWG duration (on-surface time=44.6 Vs. 45.4%, p=0.56), or the look-ahead distance (look-ahead distance=9.3 Vs. 10.7m, p=0.82). Surprisingly though, increasing walking speed caused the anticipated effect, leading participants to look longer at the walking surface (on-surface time=44.6 Vs. 68.3%, p=7×10^-5^). Increasing walking speed also led to a notable decrease in the look-ahead distance from 9.4 to 6.9m ahead (p=0.004).

**Figure 4.**
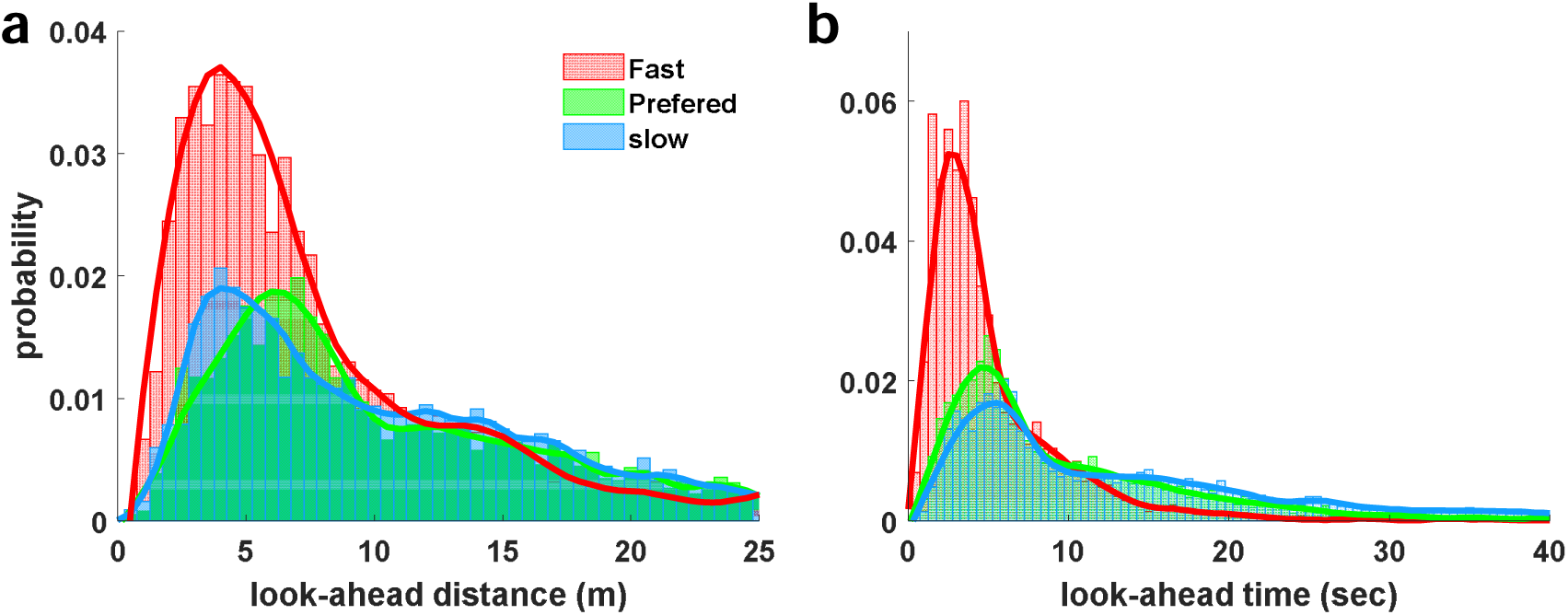
look-ahead distance (**a**) and time (**b**) in the three walking speeds: preferred, slow and fast. Slow walking speed had no effect on DWG duration, but fast walking caused an increase in duration that was comparable to the effect of stroke.

These findings are not only contradictory to what would be expected if slow walking speeds were the cause of the temporal difference between PwS and controls, they also suggest that the spatial difference is not caused by walking speed (as would be predicted from a ‘time-ahead’ strategy), but merely associated with it.

The above comparisons between PwS and controls is based on the hypothesis that the former are unstable. Nonetheless, this hypothesis was not tested and verified. To relate DWG with walking instability we wanted to test the association between the two. Therefore, in the next stage we investigated the association of DWG behavior with measures of postural and gait steadiness. To do so, postural sway and gait steadiness were evaluated in PwS (see Methods), and the correlation between these quantities and DWG was tested.

This analysis is presented in Table 3, representing a correlation matrix (Spearman correlation coefficients) of these associations. Briefly, DWG, in terms of both (increased) duration and (shorter) distance was associated with increased postural sway, and to lesser extent with increased spatiotemporal gait variability. Importantly, DWG was also associated with walking speed, but in contrast to the association observed during the walking speed manipulation, lower rather than higher speeds were associated with DWG, as would be expected from a time-ahead strategy.

Poor motor control (and the resultant instability) could be a consequence of a shift from automatic to conscious movement control [see review in [36]], rather than poor control per-se. Such a shift is often observed under conditions of elevated stress and was shown to occur in situations of increased risk of falling, and to affect gaze behavior [37, 38]. To test this possibility, we also had PwS respond to the Gait Specific Attentional Profile (G-SAP, [39]) as an estimate for their tendency for conscious movement processing (CMP) and fear of falling (Anxiety). The results of this analysis revealed that CMP was not associated with DWG while fear of falling was associated with all parameters (see Table 3). Importantly, however, PwS indeed tended to consciously control their gait, as indicated by their median score of 11 (IQR 8-13) on a 3-to-15 ordinal scale of the G-SAP.

One concern is that PwS with poor balance and gait control are more likely to use assistive devices during walking and to walk more slowly. If so, it is possible that the use of an assistive device, rather than these specific traits, affect gaze behavior.

Indeed, when we compared the steadiness of gait and posture between those who used assistive devices during walking (‘users’) and those who did not (‘non-users’), the latter were steadier and walked faster than the former (see Supplementary Materials TableS2). Next, we tested how the use of assistive devices affects gaze behavior. To do so, the gaze behavior of ‘users’ and ‘non-users’ and that of healthy controls was compared. The results of this analysis revealed that the use of an assistive device was indeed associated with a greater tendency to gaze down (‘users’ compared to ‘non-users’), but also that both ‘users’ and ‘non-users’ gazed down more than controls, indicating that the reported DWG effect is not a result of using an assistive device. (See Figure 5.)

**Figure 5.**
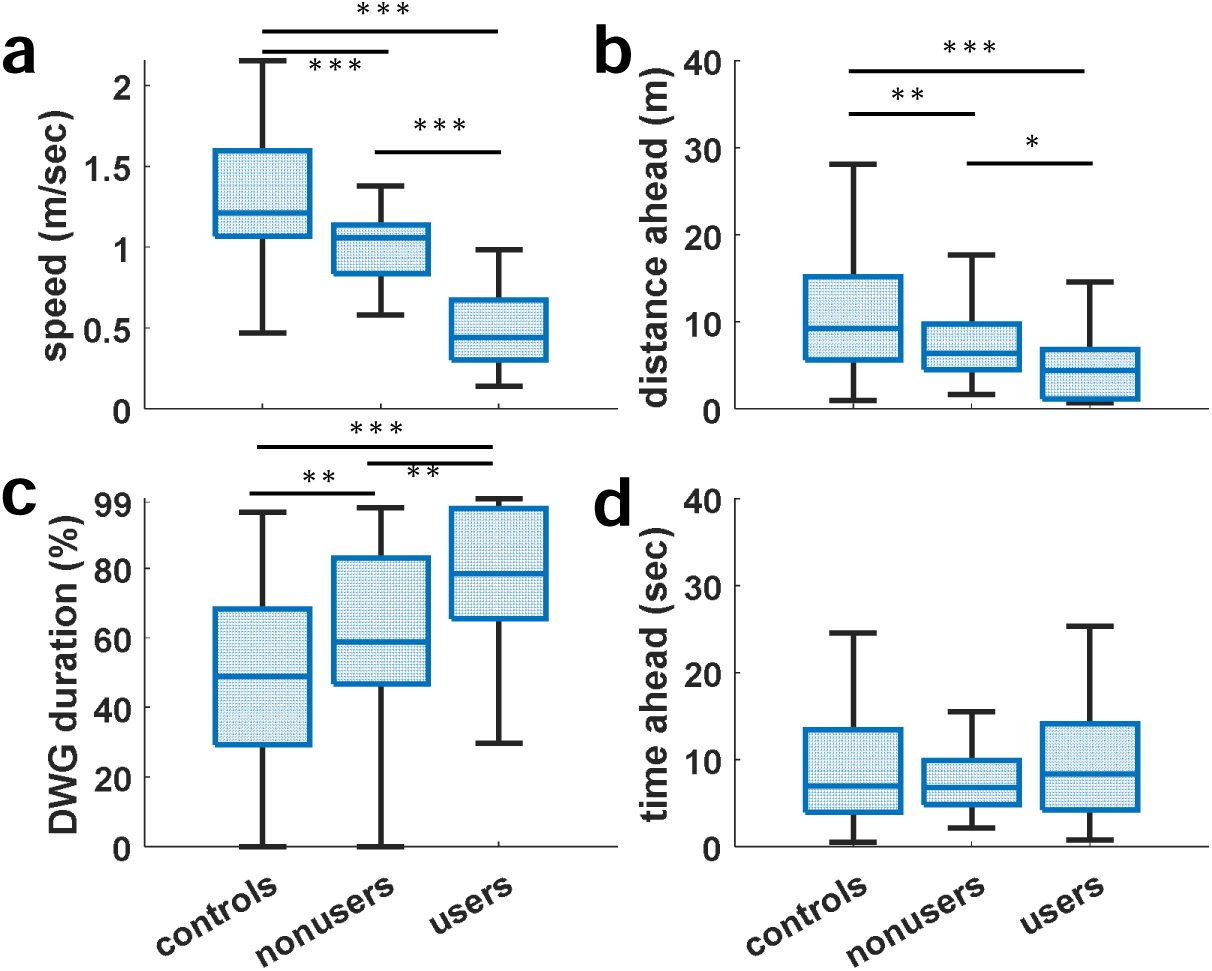
comparisons between controls, PwS who walk without, and PwS who walk with assistive devices, for (a) walking speed, (b) look-ahead distance, (c) on-surface percentage of time and (d) look-ahead time. This comparison revealed that both non-users and users walked more slowly and gazed down more than controls. The look-ahead time was not different between groups. ‘*’, ‘**’ and ‘***’ indicate p<0.05, p<0.01 and p<0.001, respectively.

## Discussion

The main objective of this investigation was to ascertain our clinical impression that PwS tend to gaze down while walking, and to provide a detailed description of the phenomenon. Clearly, our clinical impression intuitively relates DWG, in terms of distance and duration, with walking instability. The results of the current investigation do confirm this impression, but portrays a complicated picture of interactions between different components.

Spatially, PwS looked a shorter distance ahead than controls. Previous reports [40, 41] indicate that lower gaze angles are associated with the level of perceived complexity of the walking surface. While the walking surface investigated here was flat, the perceived complexity is also related to the motor abilities of the walker, so this shorter look-ahead distance might reflect such self-perceived abilities in the different groups. Yet, the most prominent spatial effect of stroke was the distinct dual-peaked, look-ahead distance distribution pattern. By contrast, healthy controls exhibited a single peaked distribution, located at a greater distance ahead than both peaks of PwS (Figure 2a). This pattern might indicate that, for PwS, the look-ahead distance distribution is comprised of two populations, within and/or between subjects. In the former case (within subject), this pattern might indicate that participants used two different gazing behaviors, possibly one that is associated with increased cognitive control, and another that is more automatic. Indeed, PwS have been reported to be more sensitive to cognitive interference [see review in [42]], and to exhibit greater cortical activity than healthy controls during walking [43, 44]. As opposed to this “within-subject” explanation, the two peaks could also represent two populations of participants. If so, what characteristic, or characteristics, distinguish between these populations?

In investigation into walking gaze behavior, it is common to use the look-ahead distance (in terms of steps ahead) as an outcome measure. A growing body of evidence indicates that precise stepping is most often planned ∼2 steps ahead [12, 13, 19, 28, 45–47]. While the current investigation did not require precise stepping, PwS reported that they actively paid attention to their stepping motion, possibly due to poor control over the paretic limb. This might be useful, for example, to minimize the energy expenditure associated with increased step-width [21, 48]. If so, the peak observed at ∼1.5m ahead might represent such conscious visuomotor control in some participants, while the other peak represents those who did not exert this effort. That said, 2, or any other number of steps, is not only a spatial measure, but also a temporal one (time taken to make the number of steps). In fact, given that, during walking, visual input is primarily used to plan future actions [8], and that acquiring and processing visual input, and planning an action are all time sensitive, time ahead might be a more reasonable outcome measure.

A time-ahead strategy implies that walking speed dictates the look-ahead distance. Therefore, the two peaks observed for PwS can simply reflect different sub- populations that walked at different speeds. Further, given that PwS walked more slowly than controls, it is possible that the spatial difference between groups simply reflects such a time-ahead strategy. To explore this possibility, we analyzed the distribution of the time-ahead over each walk. This analysis revealed a more uniform distribution between groups, with a single peak at ∼4.5sec ahead. This finding, along with the fact that walking speeds positively correlated with gaze distances, lends support to this, time-ahead possibility. In line with this observation, Matthis et al. [19] reported a similar effect observed for walking on different terrains (although they reported a peak at ∼1.5sec ahead).

While these results might indicate that humans look ahead in time, rather than space, it is important to note that a time-ahead strategy makes sense when visual input is used to plan a specific action, as was the case for [19] in which participants were required to select safe footholds. In our study, however, planning of subsequent steps was not required, so it is unclear why such a strategy would be beneficial and what exactly is planned 4.5sec ahead in time. Nonetheless, the coupling of gaze and stepping is so inherent in the control system, that it is preserved even in the absence of visual input [35]. In fact, Zubair et al. suggested that gaze-stepping coordination is not vision driven but is only modulated by the visual structure of the environment. Put simply, this speed-distance relation is not necessarily related to the acquisition and processing of visual input, but may represent a habitual, whole-body coordinated behavior.

Regardless of whether our results are vision-driven or not, it is tempting to interpret them as an indication that the two distance peaks, and the difference in gaze distance between PwS and controls, were simply a result of walking speed differences. However, the distribution of walking speeds showed no such trend (data not shown). Further, several gaze patterns were observed among participants. One of these was a gaze “parked” in space [e.g., [12, 13]]; another was fixation on some object at the end of the corridor, indicated by a gradual decrease in gaze distance and angle [e.g.,[49]]; and a third was a dual-peaked distribution (see these examples in the Supplementary Materials, Figure S3). Furthermore, when controls were asked to walk more slowly than their preferred speed, gaze distance was unaffected, indicating, at the very least, that slow speed does not cause gaze distance to decrease as would be expected from a time-ahead strategy. This might indicate that, in PwS, both walking speed and gaze distance were affected by a common source.

Temporally, PwS spent a greater percentage of time looking at the walking surface. This increase was negatively correlated with walking speed, further implicating speed in the differences between PwS and controls. Yet, while DWG distance can be directly related to walking speed, through the proposed time-ahead strategy, it is unclear why DWG duration would be affected. Furthermore, much like DWG distance, slow walking speed in controls had no effect on DWG duration. This observation further supports the notion that speed is unlikely to cause DWG (spatially or temporally), but instead that DWG and slow walking speed result from a common source. Importantly, the increase in DWG duration was not consistent across gaze distances. Specifically, a notable increase was observed at distances up to ∼7m ahead, while at greater distances, small, or no differences were observed between groups. To explore this impression, we calculated the odds-ratio between groups, throughout the range, to reveal that all distance bins were affected.

Statistical comparison of DWG duration between groups in the Short and Long look-ahead ranges provided further support for this notion. Nevertheless, we are not sure what the specific patterns observed (linear for distance and quadratic for time) mean, nor what the implications are for the relations between the odds-ratio and distance/time.

The comparison between walking gaze behavior of PwS and that of controls clearly indicates differences between these populations, but it does not indicate what drives PwS to gaze down. Our clinical impression, however, does relate DWG with instability. To investigate this relation, we tested the association of several markers of instability with the tendency among PwS to gaze down.

First, fear of falling was found to be associated with DWG. This finding is consistent with previous reports. Specifically, under conditions of induced fear of falling, the first reaction of participants was to gaze down at the immediate walking surface instead of the specified stepping targets [20, 37, 38, 50]. These authors speculated that fear of falling leads to CMP, which changes the visual requirements of the task and impairs motor-control [36]. Nonetheless, Uiga et al. [20] tested this possibility and found that gaze behavior was associated with fear of falling, but not with CMP, a finding similar to our own. This is quite remarkable given the very different methodologies used in our investigation and theirs. That said, both Uiga and our team tested the association using correlation analysis, which is not the only, and possibly not the best, way to test such an association. In fact, the median score on the CMP scale in our sample was 11 [8–13], indicating that participants were indeed trying to control their steps consciously.

Next, we investigated the association of DWG with walking instability. While induced walking instability was shown to cause DWG [1], there is limited information relating DWG with “natural” walking instability [50]. It is important to note that currently there is no consensus about the operational definition of walking stability [51], making it impossible to quantify this trait. Nonetheless, increased spatiotemporal variability of gait [see reviews in [51, 52]] and of postural sway [53–55] has been reported to differentiate between fallers and non-fallers, and to predict future falls. Therefore, regardless of their true nature, these quantities, reflecting the consistency or ‘steadiness’ of the motor output, are often regarded as indirect measures of the adequacy of the control system [56] and are used as measures of walking stability [51].

Using common measures of balance and gait ‘steadiness’, we found that an increase in all parameters of postural sway was associated with DWG distance and duration. These results possibly indicate that both gaze behavior and walking speed are affected by poor balance, either directly, or indirectly through fear of falling. Several, but not all, parameters of gait were also associated with DWG, but to a lesser extent than measures of standing postural sway. Since poor balance naturally affects gait, these properties must be separated to test which, if any, of them is associated with DWG. Here, participants held on to the handrails during their treadmill walk to increase their base of support and alleviate the balance problem (see Methods).

While this does not ensure that the increased gait variability resulted strictly from poor stepping control, it does increase the probability that poor balance was not the main cause.

As mentioned above, both balance and gait (standing sway and gait variability) were associated with DWG, but the latter to a lesser extent. However, spatiotemporal gait variability was reported to predict future falls, but not necessarily to reflect fear of falling [57]. By contrast, measures of standing sway are more likely to reflect fear of falling than the probability of falling [58]. Furthermore, walking speed is known to be associated with spatiotemporal gait variability [59]. Given that during treadmill walking participants were tested at their preferred speed, it is possible that the correlation between DWG and gait variability simply reflects the speed association of these variables. Taken together, these results draw a complicated picture of associations between DWG, walking speed, poor balance and gait control, and fear of falling.

Whether fear of falling directly drives DWG, or this is mediated through walking speed, poor control and/or CMP, as was previously suggested [20, 37, 38, 50], is unclear at this time.

In this discussion, we mention the effect or lack thereof that a forced slow walking speed had on gaze behavior. Nonetheless, the effect of a fast walking speed in healthy control subjects was remarkably similar to that of stroke. While this effect could be a consequence of the short walking distance and prolonged acceleration phase, this observation might be more meaningful. Is it possible that the behavior of PwS is driven by the fact that they walk faster than their preferred speed? Could it be simply that the biomechanics of acceleration, in which the body leans forward, is similar to that of PwS? Or is it a matter of similar visual requirements? Regardless of the answer, this effect should be explored more carefully in the future, as it is a promising avenue to understanding the phenomenon of downward gazing while walking.

Over the past years we have relied upon our clinical impression to investigate the causal relation between DWG and instability [1, 34, 60, 61]. Herein, we take a step back to ascertain our clinical impression. Overall, the current report, while confirming our clinical impression, provides only a glimpse into the altered walking gaze behavior of PwS. The picture presented here indicates that the interaction between gaze behavior, walking speed, poor balance and gait control, and psychological influences is highly complicated. There are numerous important questions that are left unanswered, such as whether DWG behavior is vision-driven? Is DWG beneficial, and if so, what are its cost and benefits? While we are continuously engaged in exploring the recovery of mobility in PwS, and particularly the use of visual input, this study only scratched the surface of the puzzle— how does the sensorimotor system react to impairment? Here we call upon the community to join our effort to explore a much-neglected domain of the motor-control of walking.

## Methods

### Participants

This study is part of an ongoing project aimed at studying the dynamics of recovery following stroke. Patients who experienced their first-ever stroke and who, prior to the stroke, had engaged in activities of daily living independently, were recruited from an inpatient rehabilitation center at Ad-Negev Rehabilitation Hospital. Prospective participants who had other neurological backgrounds, were mentally unstable, had a history of substance abuse, or were medically unstable, were excluded. Participants unable to walk, or who reported using glasses when walking and whose prescription exceeded ±3-diopter, were also excluded. Contact lenses were allowed without restrictions, but none of the participants used them.

Healthy controls (HC) were recruited from among the staff, volunteers and visitors at the hospital. All HC reported to be free of any condition that might affect walking.

Before enrollment, they were interviewed about their health to clarify what was and was not a condition that might affect walking. We recruited both younger (<40) and older controls (>50), but apart from age, no difference was found between them.

Therefore, for comparison with PwS, a single control group of participants from all ages was used.

The study was approved by the Local Ethical Committee at Adi-Negev Rehabilitation Hospital, and all participants signed a consent form prior to their enrollment.

### Procedure

All participants were fitted with a mobile eye-tracker (Pupil-Labs, Invisible, *GmbH*, Germany) and were then asked to walk at a comfortable pace along one of the corridors in the hospital (∼15m long). Their preferred walking speed was determined by using a stopwatch to measure the time it took them to walk the middle 10m of the corridor. Each participant performed two walks (back and forth one time). After this initial test, HC were also asked to walk back and forth at a slower and at a faster speed.

To ensure accurate calibration of the eye-tracker, before testing, participants were asked to fixate on a far (>10m) and a near (<3m) target. Gaze position estimates were adjusted (when necessary) according to these known target locations, using the free, online platform provided by the manufacturer (i.e., Pupil-Cloud).

#### Additional tests

Postural control was assessed in PwS, by asking subjects to stand barefoot, in a standardized narrow-base stance [61], as still as possible for 90sec.

The test was performed twice with a 2-min rest between them. To quantify sway, participants performed the test while standing on a platform with an embedded force sensor array (*Zebris Medical GmbH*, Germany). Participants who were unable to maintain balance in a narrow base stance were not tested, and these missing values were replaced with the maximal sway values observed for this group. For statistical analysis, we used the average value of the two trials.

Gait consistency/steadiness was assessed by asking participants to walk on a motorized treadmill (*Zebris Medical GmbH*, Germany) with an embedded force-sensor array for 2min at the speed recorded when walking in the corridor (mean of the two walks). All participants wore a safety harness and held on to the handrails.

In addition, PwS responded to the Gait Specific Attentional Profile (G-SAP, [39]) to assess their tendency for conscious movement processing and fear of falling (anxiety) during walking.

### Data Analysis

The gaze data-processing procedure has been previously reported [1]. Briefly, the eye-tracker provides a video recording from the walker’s perspective (using its scene camera) and a series of 2D gaze positions, projected onto this video (given in pixel dimension). Using a dedicated MATLAB script, these data were segmented to include only the walking periods. Then, an image-processing procedure was used to identify the wall lines in each frame (see Figure 1), to compute the properties of each line, and to determine the intersection point of these lines (i.e., the vanishing point). The area contained within this ‘triangle’ was defined as the future walking surface (real or imaginary, see Figure 1). Next, each gaze position was classified as either ‘On’ or ‘Off’ this surface. For ‘on-surface’ positions, the look-ahead distance was computed using simple trigonometric calculations.

These distances were binned to 0.5m intervals, and the probability of each bin was calculated (number of instances divided by the total number of samples during the entire walk). To transform look-ahead distance to look-ahead time, distances were divided by the mean walking speed and binned to 0.5sec intervals. For graphical presentation, look-ahead distance bins were limited to 25m ahead, and those of look-ahead time to 40sec ahead. These ranges contain >90% of the actual data. For consistency with [1], data was also binned into 3 distance categories: <3m, 3-10m and >10m ahead.

In addition to this data, the eye-tracker also provided the pitch angle of the eyes and head (the latter was provided by an inertial measuring unit). These can be used to calculate the gaze pitch angle (simple addition). Yet, these angles are only estimates as they are relative to the eye-tracker. Specifically, the head angle is actually the pitch angle of the eye-tracker with respect to gravity, meaning that different head anatomies will result in different angles, regardless of the head’s true angle. In the same manner, the eye angles are relative to the eye-tracker. To better represent the tendency to gaze down, we also estimated the distance (in pixel dimension) of the gaze position relative to the vanishing point (horizon, see also Figure S1). This measure is extracted from the images, using image-processing techniques, and is less sensitive to different anatomies.

As main outcome measures, we used the percentage of time participants looked onto the walking surface, and the median and median absolute deviation (MAD) of the look-ahead distance and time. In addition, we also computed the median and MAD of the eye, head, and gaze pitch angles, as well as the median and MAD of the vertical distance between the gaze position and the vanishing point. (These outcomes are not included in the main manuscript and appear in the Supplementary Materials). While look-ahead metrics were computed only for ‘on-surface’ gaze positions, the other outcome measures were computed for the entire walk, regardless of where the participant was looking.

To analyze postural sway, we processed the center of pressure time series data as described in [34, 60, 61]. As outcome measures for postural control, we used the traditional sway metrics, i.e., sway range in the anterior-posterior (AP) and the mediolateral (ML) directions, sway area and sway velocity. We also used the short-term diffusion coefficients for the 1-D (ML and AP) and 2-D sway, as derived from stabilogram diffusion analysis [62].

As a quantitative measure of gait steadiness we used the standard deviation of step time, length, width, and velocity. The procedure used to process and analyze the center-of-pressure time-series data, extracted from the force-sensor array, are described in detail elsewhere [59].

### Statistical analysis

All tests were performed using the statistical package for social sciences (SPSS, V.29 Armonk, NY). To compare gaze behavior between PwS and controls, we used linear mixed-effects models. In several cases, PwS were tested on more the one occasion (2-6 months apart). In the models, testing sessions (rather than the subject) were used as the random effect, as we were not interested in the within-subject effect. (The random effect was used to control for the two walks in each testing session.) In the models we used the group/walking-speed category (slow, preferred, and fast) as the fixed effects. For post-hoc pairwise comparisons (when required), we used a paired samples t-test. Other comparisons between groups (appearing in the Supplementary Materials) were preformed using the Mann-Whitney U test. To test the relation between the odds-ratios and gaze distance/time, we used linear models. For correlation analysis, we used the Spearman correlation coefficient, after averaging the gaze behavior in the two walks. Here, we decided not to set a threshold for significance, but instead to report the probability for a type-1 error as accurately as possible. We leave it up to the reader to decide if results are meaningful or not. Note that SPSS has a limit of 15 decimal places, so p=0.00 in the text indicates that p-values are smaller than 1×10^-15^.

## Data Availability

Data will be made available upon request. Once published in a peer-reviewed journal, data will be made publicly available.

## Declarations

### Funding

The author(s) disclosed receipt of the following financial support for the research, authorship, and/or publication of this article: this research was supported by the Israeli Science Foundation (grant 1244/22 to LS) and the Lillian and David E. Feldman Research fund.

### Declaration of Conflicting Interests

The authors declare that the research was conducted in the absence of any commercial or financial relationships that could be construed as a potential conflict of interest.

### Ethics approval

The study was approved by the Ethics Committee at Adi-Negev Nahalat-Eran Rehabilitation hospital, Ofakim, Israel (ADINEGEV-2023_106), and conformed to the standards set by the Declaration of Helsinki. All participants provided written consent prior to their enrolment.

## Notes

### Competing Interest Statement

The authors have declared no competing interest.

