## Supplementary Materials for "Walking Gaze Behavior After a Stroke: More Than Meets the Eye"

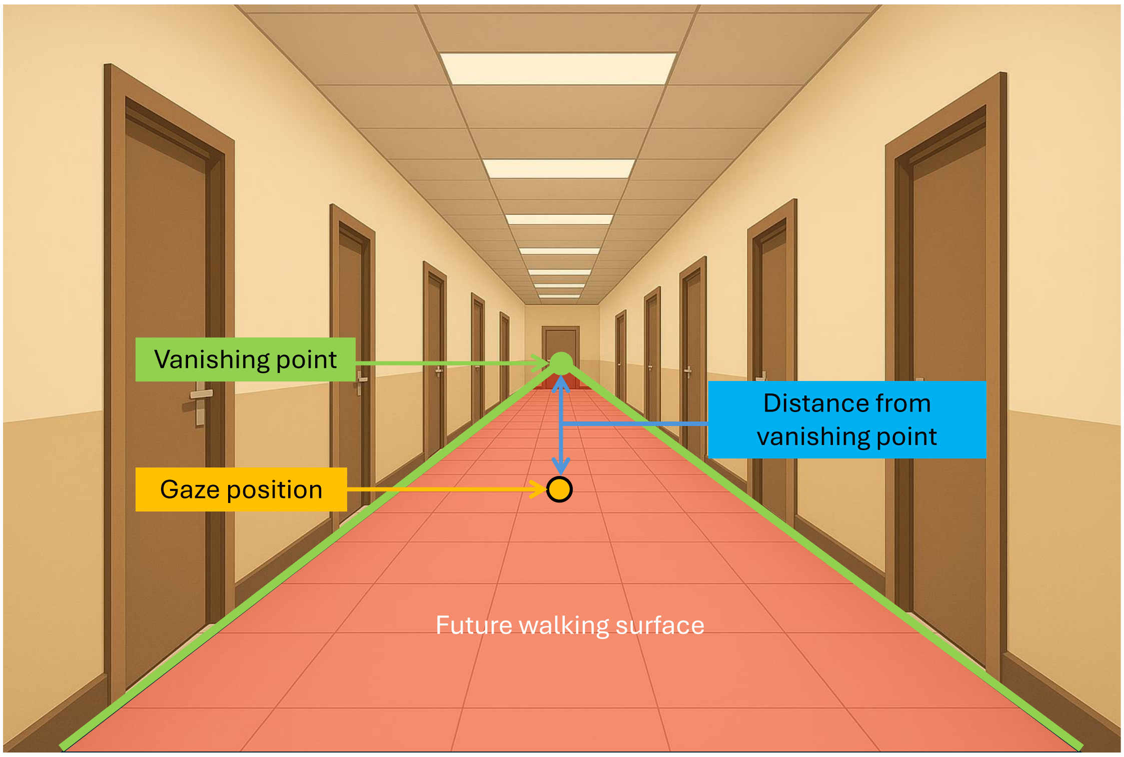


*Figure S1-Illustration of the experimental setup [from a previous study. Participants, wearing a mobile eye-tracker, walked back and forth along a corridor. Using image processing, we identified the floor-lines (highlighted in green) and their intersection point—the vanishing point (the green circle) in each frame. The area contained within this "triangle" (shaded in red) is the future walking surface. The vertical gaze position is the distance (in pixels) between the Y-value of the vanishing point and that of the gaze position (orange circle). Values are reported as percentage of image size. Gaze positions projected onto the walking surface (shaded red) are "on surface" and are reported as percentage of all gaze positions during the entire walk (DWG duration). Each "on-surface" position is located at a unique distance from the observer, reported in meters (DWG distance). For eye, head, and gaze (sum of eye and head) angles, center and spread values were computed for the entire walk (regardless of whether gaze was directed onto the walking surface). For analysis, we used the median and the median-absolute deviation (MAD) as center and spread measures of gaze parameters in each walk.*

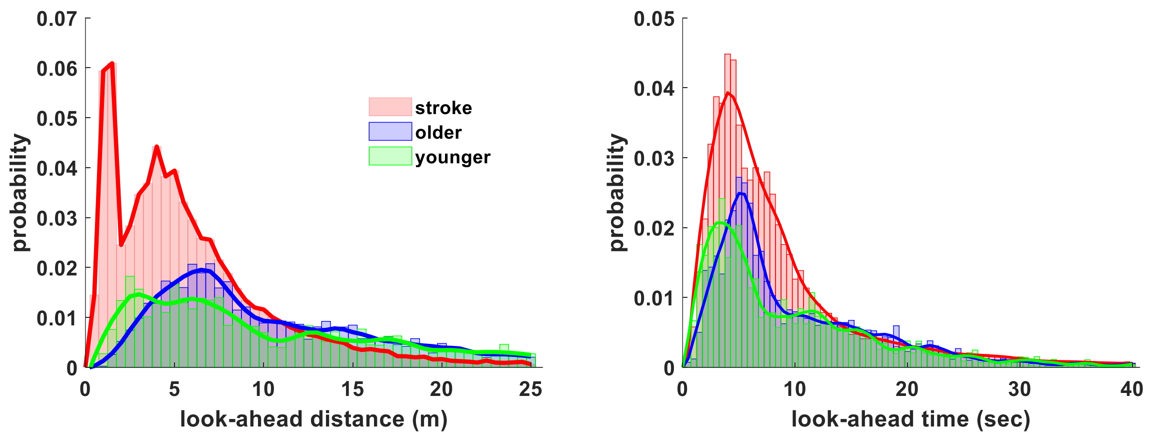


*Figure S2- The distribution of the look-ahead distance (left) and time (right) in healthy younger and older adults, and in persons with stroke. Neither look-ahead distance, nor look-ahead time were different between the younger and older healthy adults, so these groups were combined into a single control group.*

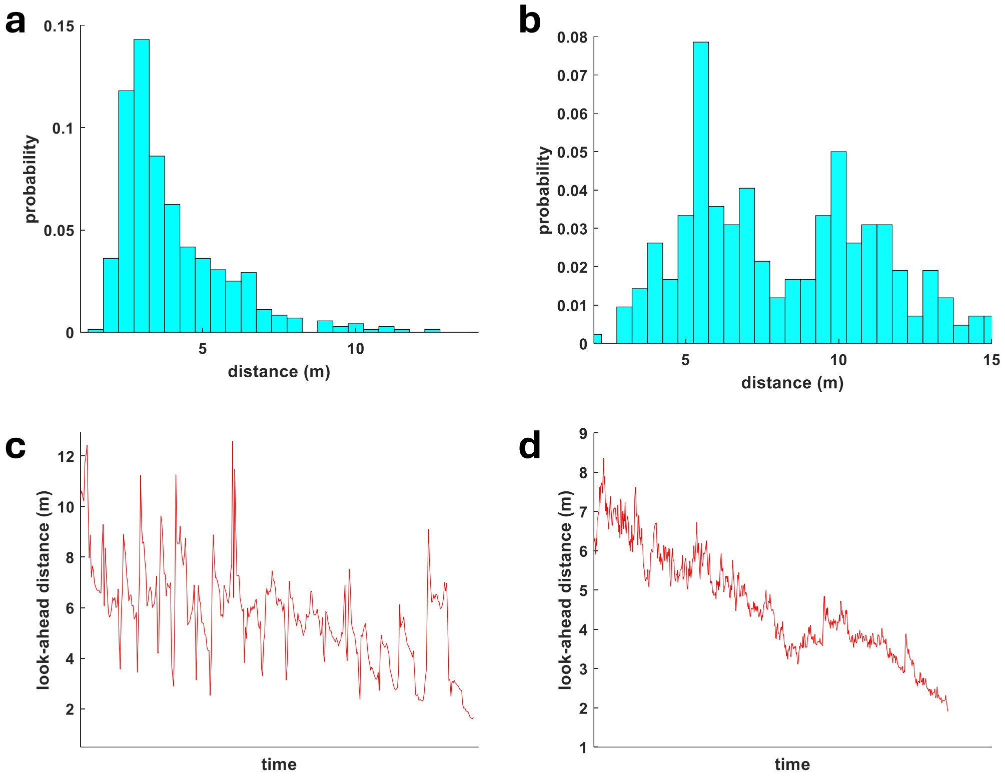


*Figure S3- Examples of several walking gaze behaviors. Pannel **a** presents a Gamma-like distribution of the look-ahead distance (most participants). Pannel **b** presents a dual-peak distribution. Pannel **c** presents a gaze that seems to be synchronous with the stepping motion. Pannel **d** presents a pattern of decreasing gaze distance over time, indicating that the participant was fixating on some object at the end of the corridor.*

Table S1-Comparison of the vertical eye, head, and gaze (sum of eye and head) angles between groups. Angles are relative to the horizontal plane, such that positive angles are above this plane, and negative angles are below it. From the descriptive statistics, it would appear as if participants generally looked up (above the plane) and not down. This impression, however, is wrong because for these angles to be true, the plane of the eye-tracker and that of the face would have needed to be parallel, which is impossible given the different facial anatomies. To better represent the vertical gaze angle, we used the vertical distance of the gaze position from the vanishing point (see Figure S1). This estimate is directly extracted from the image and is therefore less biased. Indeed, the median gaze position in both groups indicate downward gaze angles (relative to the horizontal plane).

| **Median [IQR]** | HC | Stroke | p-value |
|---|---|---|---|
| Eye angle° | 4.1 [-1.3-7] | 4.5 [1.3-10.4] | =0.21 |
| Eye angle MAD° | 2.1[1.5-3.4] | 3.1 [2.2-4.2] | =0.003 |
| Head angle° | 6.8 [2.7-12.9] | -4.9 [-11.3-1.7] | <0.001 |
| Head angle MAD° | 2.1 [1.7-2.9] | 2.7 [2.2-3.5] | =0.01 |
| Gaze angle° | 11 [8.4-12.8] | 3.4 [-4.4-6.6] | <0.001 |
| Vertical position % | -6.3 [-10.3-(-3.5)] | -17.6 [-29.8-(-11.4)] | =0.002 |
| Vertical position MAD % | 47.3 [43.7-54.2] | 48.6 [40.5-54.9] | =0.83 |

**For simplicity, we used the Mann-Whitney U test to compare between groups.*

Table S2- standing and walking steadiness was compared between PwS who use assistive devices (of any kind) for walking and those who do not. Comparisons of their level of anxiety (fear of falling) and tendency to consciously control their walking are also presented. All comparisons are based on Mann-Whitney U test.

| | None-users | Users | p-value |
|---|---|---|---|
| | Standing sway (median [IQR]) | | |
| ML-Range *mm* | 31.3 [24.2-40.1] | 46.8 [32.5-95.2] | =0.00 |
| AP-Range *mm* | 27.9 [23.1-33.2] | 48.2 [31.3-70.3] | =0.00 |
| Velocity *mm/sec* | 15.0 [12.7-18.7] | 28.5 [18.2-42.1] | =0.00 |
| Area $mm^2$ | 567 [369-857] | 1657 [658-3794] | =0.00 |
| Drs $mm^2/sec$ | 29.5 [18.7-44.1] | 101.9 [39.1-386.0] | =0.00 |
| Dys $mm^2/sec$ | 11.8 [7.9-16.9] | 45.5 [19.9-111.7] | =0.00 |
| Dxs $mm^2/sec$ | 17.4 [11.7-25.6] | 51.6 [22.5-331.3] | =0.00 |
| | Walking speed and step variability (median [IQR]) | | |
| Walking speed *m/sec* | 1.05 [0.85-1.13] | 0.44 [0.30-0.67] | =0.00 |
| Time SD $sec(\times 10^{-2})$ | 2.6 [2.1-4.0] | 5.1 [3.8-7.6] | =0.00 |
| Length SD *cm* | 4.0 [2.4-5.4] | 4.2 [3.8-6.7] | =0.16 |
| Width SD *cm* | 12.3 [10.1-15.1] | 14.3 [11.9-23.9] | =0.03 |
| Velocity SD *cm /sec* | 5.4 [3.2-7.4] | 5.4 [3.4-8.9] | =0.74 |
| | Psychological factors (median [IQR]) | | |
| Anxiety | 4 [3-7.5] | 7 [6-9] | =0.03 |
| CMP | 11 [6.5-13] | 11 [9-14] | =0.35 |
